# Initial experiences with *Mycobacterium tuberculosis* DNA extraction for downstream Deeplex Myc-TB targeted deep sequencing in a high burden setting

**DOI:** 10.1101/2023.11.22.23296677

**Authors:** Jason D Limberis, Alina Nalyvayko, Janré Steyn, Jennifer Williams, Melanie Grobbelaar, Robin M Warren, John Z Metcalfe

## Abstract

The propensity for *M. tuberculosis* to develop resistance and the lack of clinical tools for the rapid determination of such resistance has long significantly complicated tuberculosis (TB) therapeutics. Targeted next-generation sequencing (NGS) has improved our understanding of the genetic basis and identification of drug-resistant TB. However, to achieve accurate results reliable enough for clinical implementation, high-quality *M. tuberculosis* DNA must be extracted from patient-derived samples within high burden routine laboratory workflows. In advance of a large cluster RCT in the Western Cape of South Africa evaluating the Deeplex Myc-TB targeted NGS assay (GenoScreen; Lille, France), we sought to compare DNA extraction methods for both early MGIT culture-positive samples and processed patient sputum. Given the lack of reference standard method, we assessed a representative set of DNA extraction protocols, including the GenoScreen-recommended method, in parallel in South Africa and at UC San Francisco. Our findings provide preliminary insights into an optimal DNA extraction method for the utilization of Deeplex Myc-TB in routine laboratory settings and can inform future experiments evaluating newer generation assays.

## Introduction

Tuberculosis (TB) is a significant public health concern causing 1.5 million deaths annually^1^. Drug-resistant *M. tuberculosis* strains lead to prolonged illness, increased healthcare costs, and higher mortality rates^1^. Identifying and treating drug-resistant TB rapidly is crucial to reduce morbidity and mortality and prevent onward transmission.

Targeted next-generation sequencing (NGS) has revolutionized the understanding of the genetic basis and identification of drug-resistant TB. However, to achieve accurate results reliable enough for clinical implementation, it is essential to extract high-quality *Mycobacterium tuberculosis* DNA from patient-derived samples. Therefore, the choice of DNA extraction method, typically soft-pedalled in targeted NGS publications or concealed through the exclusive use of culture isolates or high bacillary grade specimens, is critical for real-world downstream molecular assays.

*M. tuberculosis* is known for its unique cell wall, which is tough and contains high levels of mycolic acids and other lipids that make it difficult to lyse. There is substantial variation in the published lysis (e.g., sonication, chemical, heat, and bead-beating) and DNA extraction methods (e.g., phenol-chloroform extraction, ethanol precipitation, and column and bead-based methods) for *M. tuberculosis*, which result in differences in DNA yield, purity, and quality^2–17^. In addition, the same method is seldom used by more than one group, and the measure of method success varies. Thus, the “best” method for DNA extraction remains unstandardized and depends heavily on the downstream application (e.g., resolving *M. tuberculosis* structural variants directly from sputum using long read sequencing will require a higher quality DNA extraction and a higher fidelity polymerase for amplification and sequencing than Xpert Ultra targeting a small region of *rpoB*) using a highfidelity polymerase for amplification and sequencing or a robust polymerase for detection of a gene, or requiring large fragments for PCR of entire genes). Adding complexity, the yield of an individual extraction method depends on the type of sample, bacterial load, and the presence of inhibitors and co-extracted nontarget DNA, as well as variable reliance on laboratory technician experience and operator variability (e.g., differences in pipetting technique). This context highlights the need for an optimized, standardized protocol for *M. tuberculosis* DNA extraction for specific applications to advance the consistency of results across laboratories, which would reinforce the reliability and comparability of results.

In this study, we present our initial experience assessing a representative set of *M. tuberculosis* DNA extraction methods utilizing *M. tuberculosis* H37Rv and spiked human sputum at the University of California San Francisco (UCSF); a select subset of DNA extraction methods were then performed on early MGIT culture-positive samples and clinical sputum samples in a high TB burden setting in South Africa. Extracted *M. tuberculosis* DNA was used for the Deeplex Myc-TB (GenoScreen, France) targeted NGS assay, a commercial 24-plex PCR assay targeting 18 genes associated with first- and second-line drug resistance. Our measure of success for the DNA extraction methods was sufficient sequence data generated from the Deeplex Myc-TB assay and library preparation for the Deeplex Web Application to call resistance (and wildtype) mutations for the included targets (but the specific mutations were not considered when assessing the best DNA extraction method).

## Methods

### Procedures at the University of California, San Francisco

#### Bacterial culture

*M. tuberculosis* H37Rv mc^2^ 7901 was cultured at 37°C with shaking in Middlebrook 7H9 media supplemented with ADC (albumin, dextrose, and catalase) and pantothenate, L-leucine, L-arginine, and L-methionine at 24, 50, 200, 50 mg/litre to mid-log growth. The culture was centrifuged, and the bacterial pellet was washed with phosphate-buffered saline (PBS) buffer before resuspension in PBS. The culture was passed through a 30-gauge needle 10 times to reduce clumps and was then diluted to 8.4x10^6^ cells (corresponding to ∼2ml of positive mycobacteria growth indicator tubes [MGIT] culture at 200 growth units [GU]) per 50μl which were stored at -80°C until use. Dilutions were plated on Middlebrook 7H10 supplemented with glycerol and ACD, pre- and post-freezing to determine the number of cells present.

#### Spike-in sputum processing

Sputum from several individuals with non-tuberculosis respiratory illnesses was obtained from Discovery Life Sciences, Inc. (USA), pooled, and 1ml aliquots were made and stored at -80°C until use. For spike-in experiments, the desired number of bacteria in a 50µl aliquot was added to thawed sputum, vortexed for 30 seconds and left to stand at room temperature for 10 minutes. After adding four volumes of 100mM Dithiothreitol (DTT), the sample was vortexed and left to incubate at room temperature for 15 minutes. The resulting liquified sputum was centrifuged, the supernatant discarded, and the pellet containing the bacteria and debris, was resuspended in the appropriate buffer for the downstream extraction method.

#### DNA extraction methods

##### M1: Bead beating and M2: heat lysis

Pellets were resuspended in 200µl of either TE buffer or a buffer consisting of 100mM NaCl, 10mM Tris-HCl (pH 8.3), 1mM EDTA (pH 9.0), and 1% Triton X-100. Samples were heated in a dry heatblock to 90°C for 5 minutes with a heated lid set to 105°C. For samples undergoing bead beating, 125µl additional buffer was added, and bead beating was done in 1.5ml screw cap tubes (Sarstedt, Germany) using 0.1mm Zirconium beads for three rounds of 45 seconds at 6.5m/s using the Fastprep 24 (MPbio, USA), with 30 seconds resting at room temperature between cycles. A full protocol is available on protocols.io^18^.

##### M3: GenoScreen-recommended method (user manual, v5)

The Deeplex Myco-TB user manual method was carried out as recommended by the supplier^19^. Briefly, the pellet was resuspended in 250μl of 10mM TE buffer and incubated at 95°C for 15 minutes. The solution was then transferred to a 1.5 ml microtube containing ∼0.5 g of 0.1mm zirconium beads and vortexed for 30 seconds. The sample was incubated at -20°C for 30 minutes, thawed, and the supernatant was transferred to a new tube. One microliter of glycogen, 0.1 volume of 3M sodium acetate (pH 5.2), and three-volume ice-cold ethanol were added and incubated at -20°C for 10 min, followed by centrifugation at 15,000 relative centrifugal force (RCF) for 20 minutes. The supernatant was discarded, and 600μl ice-cold 70% ethanol was added before centrifugation at 15,000RCF for 5 minutes. The supernatant was discarded and the pellet air dried for 15 minutes before resuspension in 20μl of water.

##### M4: GenoLyse kit

The GenoLyse kit was carried out as recommended by the supplier^20^. Briefly, the sample was resuspended in 100µl Lysis Buffer and incubated for 5 minutes at 95°C. One-hundred microliters of Neutralization Buffer was then added and the samples were centrifuged at full speed for 5 minutes. The ∼200µl supernatant was transferred to a new tube.

##### M5: NucleoMAG kit

The NucleoMag Pathogen kit (Macherey-Nagel, Germany) for viral and bacterial RNA/DNA from clinical samples was carried out as recommended by the supplier^21^. Briefly, samples were lysed with Proteinase K and NPL1 buffer at 56°C for 1 hour. NPB2 and B-Beads we added. The beads were then separated on a magnet, the supernatant was removed, and the beads were washed with NPW3 buffer, NPW4 buffer, and finally 80% ethanol before the DNA was eluted.

#### b: Bead cleanup

1.2X volumes of AMPure XP beads (BD Biosciences, USA) were added to the sample and mixed by pipetting 10 times. The tubes were placed on a magnetic rack, 200μl of fresh 70% ethanol was added and aspirated after 30 seconds. This was repeated. The beads were allowed to dry briefly, resuspended in 20 low EDTA TE buffer, and incubated for 5 minutes. The tubes were placed on a magnetic rack and the supernatant was transferred to a new tube.

#### e: Ethanol precipitation

Two microliters of Glycoblue (Ambion, USA) was added as a co-precipitant to the sample, followed by 20μl 3M sodium acetate. The sample was mixed and 450μl ice-cold 95% ethanol added and vortex. The solution was incubated at -20°C for 1 hour then centrifuged at ≥17000RCF for 15 minutes at 4°C. The supernatant was discarded, leaving the blue pellet intact, which was washed once with 1ml ice-cold 70% ethanol and allowed to air dry for ∼5 minutes. The pellet was resuspended in 20μl low EDTA TE buffer.

#### Deeplex PCR and library preparation

Deeplex PCR was carried out as described in the User Manual (December 2022; V5) using a BioRad C1000 thermocycler; however, the ramp rate used was 2.3°C/sec (block rate 3.3°C/sec) for the sample as opposed to the 3.0°C/sec for the sample block described in the User Manual (i.e., the thermocycler standard ramp rate was mistakenly not adjusted to that recommended in the User Manual, an issue analogous to that which has frequently occurred with the Hain LPA^22^). Sequencing libraries were prepared using the Nextera XT DNA Library Preparation kit (Illumina, USA) according to Reference Guide (Document #15031942 v05, May 2019) with the following Deeplex User Manual-recommended modifications: “Use 5 μl of input DNA at 0.2 ng/μl (i.e. a total of 1 ng) for each library; At Clean-Up Libraries Step 4: Use 30μl Agencourt AMPure; Step 13: 26.5μl of RSB (resuspension buffer) instead of 52.5μl”. Libraries were quantified using the Qubit (AppliedBiosystems, USA) DNA high sensitivity kit. Libraries were spiked with 1% PhiX control and 150bp pair-end sequencing done on a MiSeq (Illumina, USA). Sequence data is available on the NIH Sequence Read Archive (SRA accession number PRJNA974051).

#### Quantitative PCR

Quantitative PCR (qPCR) reactions were done using Luna Universal Probe One-Step Reaction Mix (NEB, USA) in 10 µl reactions with TaqMan MGB Probes targeting the 16s (NED), *atpE* (JUN), *rpoB* (VIC) genes. The cycle parameters were as follows: initial denaturation at 95°C for 1 minute, then 40 cycles of 95°C for 15 seconds and 60°C for 30 seconds.

**Table 1.**
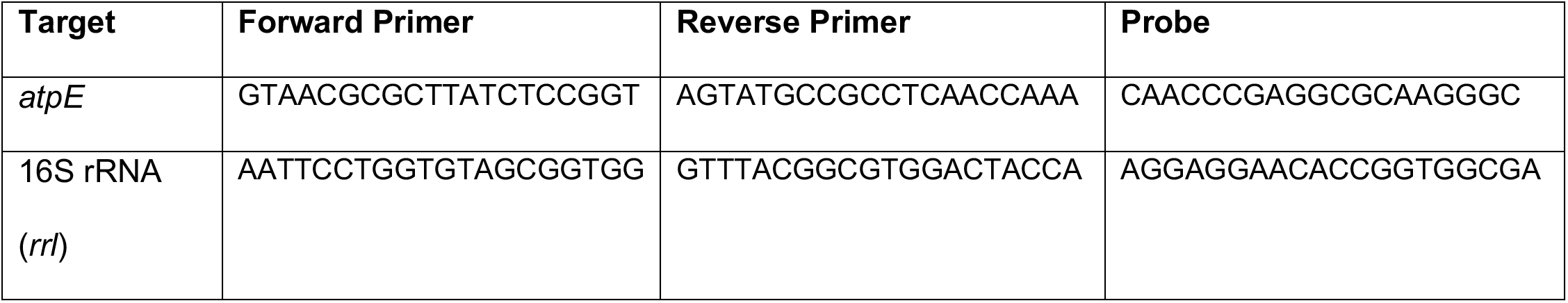
Primer sequences used in this study.

#### Bioinformatics

Reads were uploaded to the Deeplex Web Application^23^ and analysed using the “Deeplex Myc-TB V3.0.1 - Extended catalogue”. Secondary analyses were done by aligning reads first to the *M. tuberculosis* H37Rv genome (NC_000962.3) using bowtie2 (v2.4.5) with unaligned reads outputted. The unaligned reads were aligned using bowtie2 to the human genome (GENCODE 43). Bedtools (v2.30.0) was used to get per-site genome coverage, and further analyses were done in R software (v4.2).

### Procedures at Stellenbosch University, South Africa

#### MGIT bacterial culture

MGIT tubes were inoculated with patient isolates and incubated at 37°C until positivity (200GU). The MIGIT tubes were heated directly with 7.5ml of the original media for 1 hour at 80°C according to the standard operating procedures of the South African National Health Laboratory Services (NHLS) facility. We pooled 7.5ml of MGIT culture with a mean score of 1290±673 GU from 8 de-identified patient samples, which approximates to 8.46 CFU/ml. Patients had a mix of drug sensitive and drug resistant TB as determined by Xpert MTB/RIF Ultra. To extract DNA, the eight decontaminated MGIT tubes were pooled, split into twelve tubes with 5 mililiters per tube, centrifuged at 3000RCF for 10 minutes, and resuspended in the appropriate buffer according to each DNA extraction protocol.

#### Clinical sputum processing

Sputum samples were obtained from recently diagnosed TB patients and Auramine-O microscopy was done to assess bacterial load. The samples were scored as having either high or low bacterial burden using Auramine-O microscopy. The sputum samples were then processed using Mycoprep (BD, USA) according to the manufacturer’s instructions, resuspended in 350μl of the Triton X based buffer, and heated for 1 hour at 80°C. Sputum samples from each group were pooled, mixed and redistributed equally for subsequent DNA extraction.

#### DNA extraction methods (and deviations from those performed at the University of California, San Francisco)

DNA extractions were performed at Stellenbosch University (Western Cape, South Africa) by or under the supervision of the same person as at UCSF (*co-author, AN*); however, due to equipment or reagent availability protocol alterations included the following:

##### M1: Bead beating and M2: heat lysis

Bead beating was done using a Hybaid Ribolyser at the same speed and time settings as at UCSF the Fastprep 24 (MPbio, USA). Snap lid DNA low bind tubes were used instead of screwcap tubes, resulting in some sample being lost during processing. Heat lysis was done for 15min at 90°C in a dry heat block without a heated lid or agitation (in addition to heating the cultures for 1 hour at 80°C).

##### M5: NucleoMAG kit

Proteinase K was increased from 15min to 30min, RNA carrier was omitted, and the bead drying was increased from 10min to ∼30min.

There were no deviations for Method 3 (*GenoScreen-recommended method*) or Method 4 (*GenoLyse kit*), nor for bead clean-up or ethanol precipitation steps.

#### Deeplex PCR and library preparation

Deeplex PCR was performed with a block ramp rate of 3°C/sec (approximately 2.3°C/sec sample ramp rate as recommended in the user manual) using an Applied Biosystems MiniAmp Plus thermocycler. Sequencing libraries were prepared using the Nextera DNA Flex Library Prep Kit (Illumina, USA) and not the Nextera XT DNA Library Preparation kit as was used at UCSF. Libraries were prepared according to Reference Guide (Document #1000000025416 v07, May 2019) with the following modifications outlined in the Deeplex user manual: “Use 5 μl of input DNA at 0.2 ng/μl (i.e. a total of 1ng) for each library; At “Amplify Tagmented DNA” follow the procedure for 1-9 ng of input DNA”. As at UCSF, sequencing libraries were quantified using the Qubit (AppliedBiosystems, USA) DNA high sensitivity kit; spiked with 1% PhiX control and 150bp pair-end sequencing done on a MiniSeq (Illumina, USA); and is available on the NIH Sequence Read Archive (SRA accession number PRJNA974051).

#### Ethical approvals

This study was approved by the Stellenbosch Health Research Ethics Committee N21/09/093 (Project ID: 22873) and the UCSF Institutional Review Board approval for this (IRB study #: 20-33172).

## Results

### Assessment of DNA extraction protocols at UCSF

We executed several representative DNA extraction protocols utilizing *M. tuberculosis* culture isolates and sputum samples, chosen for their ability to be pragmatically implemented in routine laboratories, their likelihood of success, and their representation in the literature (**Figure 1**); the method outlined in the Deeplex User Manual (v5, December 2022) was done in parallel as a reference comparator. We performed a quantitative PCR utilizing several *M. tuberculosis*-specific targets on all DNA-extracted samples to assess starting material for Deeplex Myc-TB library preparation (**Figure 2**). The NucleoMAG extraction kit failed on two independent replicates. All other extraction methods yielded sufficient library for sequencing. All samples including those in which sequencing profiles were indeterminate had similar sequencing data availability, with >7x10^5^ reads for culture isolates, sputum spiked with 10,000 bacilli (“AFB scanty”), and sputum spiked with 50,000 bacilli (“AFB 1+”).

**FIGURE 1:**
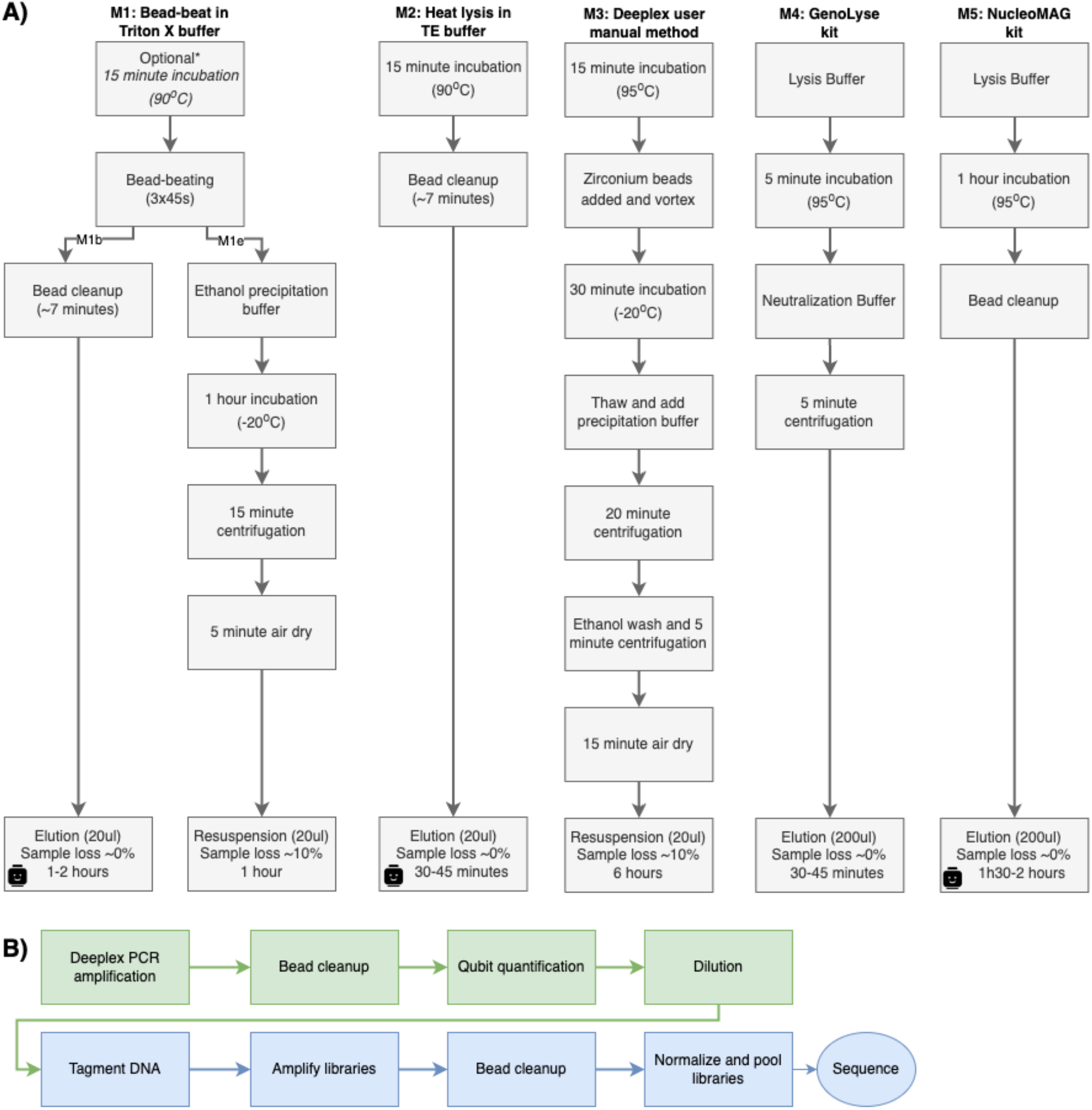
Workflow of A) DNA extraction methods used in this study and B) The Deeplex Myc-TB targeted sequencing assay workflow. Sample loss indicates the volume of the initial sample not taken forward to the end of the protocol. The Lego head indicates an easily automatable workflow. *Optional heat kill allows for removing culture sample from the biosafety level 3 laboratory.

**FIGURE 2:**
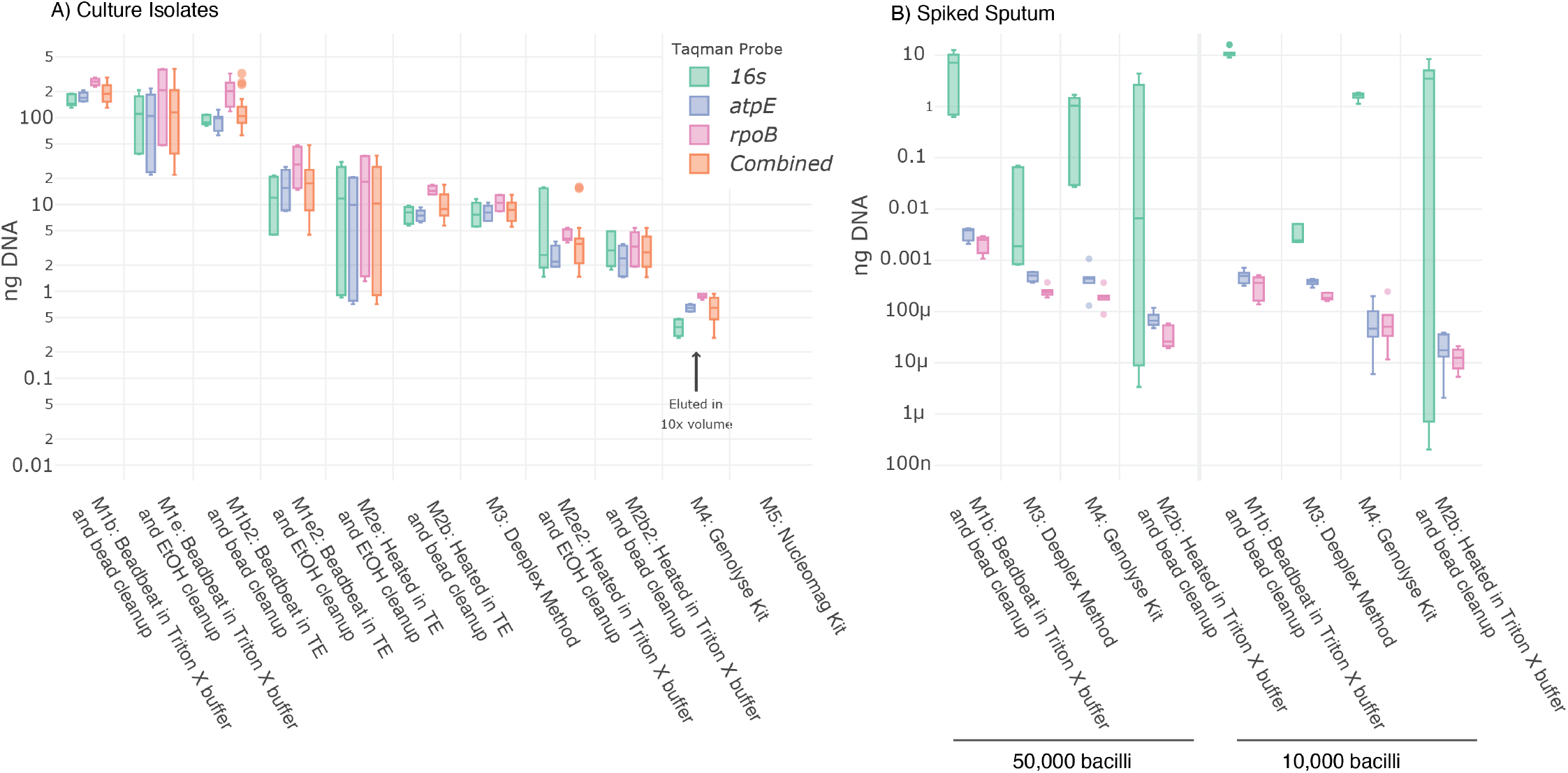
Quantifying *M. tuberculosis* DNA using three TaqMan probes in a single reaction shows successful detection from all extraction methods (Figure 1) except the NucleoMAG kit Method. A) DNA extraction from culture isolate equivalent to 2ml of MGIT positive culture at 200GU (8.4x10^6^ CFU). B) Sputum from non-tuberculosis clinical sputum pooled from patients without TB spiked with 10,000 and 50,000 CFU. The performance of the different probe targets varies markedly in these samples. Samples were done in triplicate and qPCR in technical triplicates.

#### Sequencing of culture isolates

All methods yielded successful sequencing of most drug resistance conferring regions (**Figure 3, upper portion**). However, there were erroneous resistance calls for fluoroquinolones and ethionamide for three of five methods, and several regions were not covered adequately for resistance calls in the heat lysis method.

**FIGURE 3:**
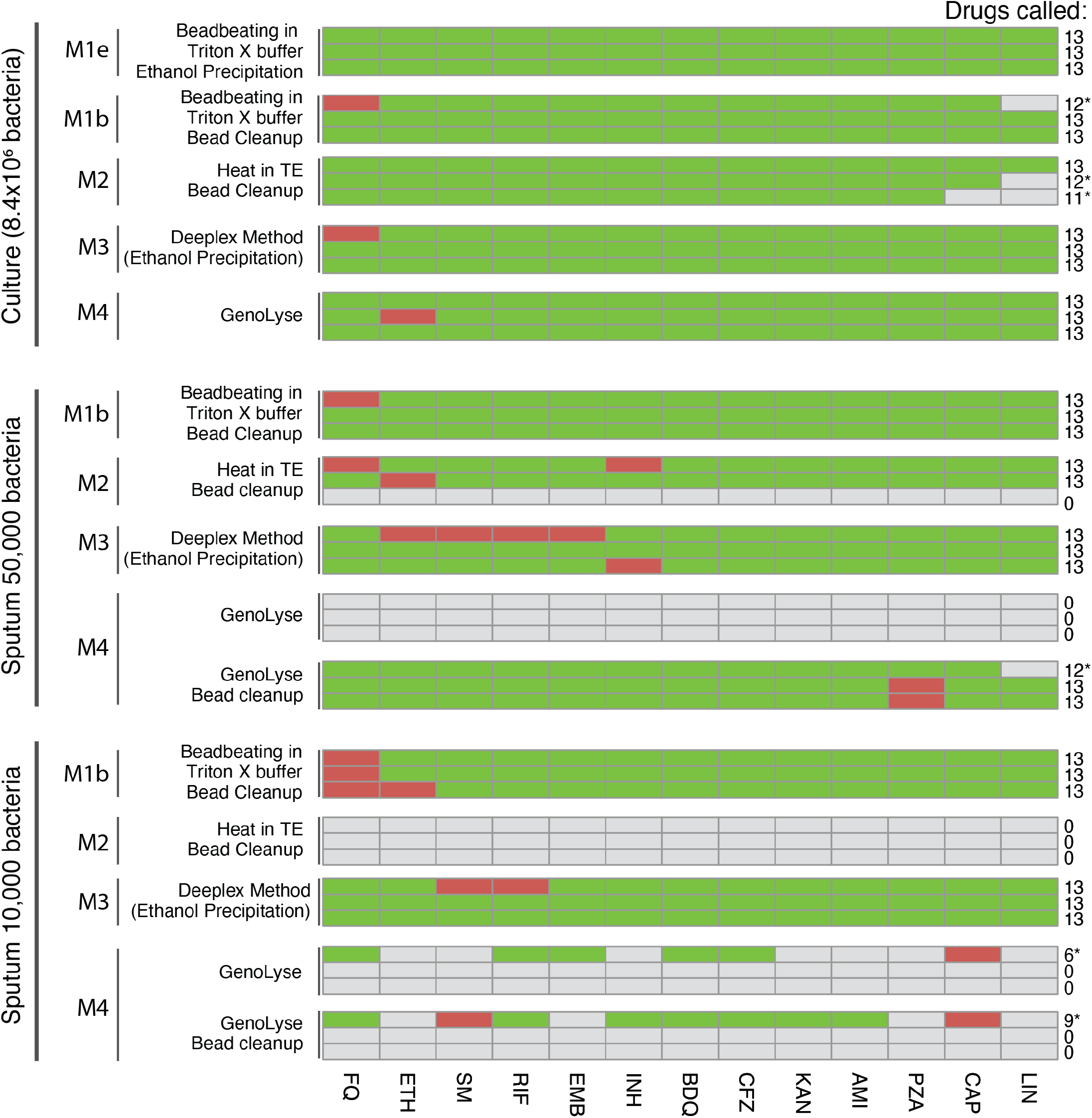
Heat map demonstrating Deeplex drug resistance call variability for the pan-susceptible laboratory strain *M. tuberculosis* H37Rv using different DNA extraction methods. The heatmap is derived from the GenoScreen Deeplex Web Application analysis. Each method was performed in triplicate. A green block indicates that a gene was correctly identified as susceptible; a red block indicates the target was amplified and sequenced but incorrectly identified compared to the H37Rv reference sequence for the drug noted on the x-axis; and the grey block indicates no or insufficient sequence data, most likely due to no amplification product formed. Ten thousand and 50,000 bacteria correspond to a Ziehl–Neelsen smear grade of scanty and 1+, respectively. *Low coverage was reported for all targets, but resistance was still called. FQ=fluoroquinolones, ETH=ethionamide, SM=streptomycin, RIF=rifampicin, EMB=ethambutol, INH=isoniazid, BDQ=bedaquiline, CFZ=clofazimine, KAN=kanamycin, AMI=amikacin, PZA=pyrazinamide, CAP=capreomycin, LIN=linezolid, MGIT=Mycobacteria Growth Indicator Tube.

#### Sequencing of sputum samples

Only bead beating in Trition X buffer with a bead cleanup and the Deeplex manual method that uses ethanol precipitation gave resistance profiles for all samples (**Figure 3, lower portion**). However, across each stratum of bacterial load, there were false positive resistance calls for one or more drugs for all methods. The GenoLyse extracted DNA showed unexpected sequencing results: *M. bovis* DNA was detected, incorrect mutations were called, and there was a high failure rate, even with concentrated samples.

#### Off-target sequencing

Samples extracted from culture using the heat lysis in TE buffer with bead cleanup and bead-beating in Triton X buffer with bead cleanup had many off-target sequences that aligned to the *M. tuberculosis* genome. We did not have access to the Deeplex Myc-TB primer sequences, so we could not accurately determine which positions were intentional and arose from amplicons; however, positions covered in the positive control accounted for 9 (IQR 7, 16) and 6 (IQR 4, 9) of the reads for the heat lysis and Triton X buffer with bead cleanup methods, respectively. Twenty-five samples, all from spiked sputum, had a median alignment to the *M. tuberculosis* H37Rv genome of only 23% (IQR 4, 39%). Most of the sequenced DNA aligned to the human genome (89%±17%), and therefore likely originated from the sputum samples.

#### Strain typing

The Deeplex Myc-TB assay contains several PCR targets for delineating spoligotypes and lineage-specific SNPs. All samples contained *M. tuberculosis* H37Rv mc^2^ 7901, which is a lineage 4 strain. However, a lineage-specific SNP was only detected in one sample (1/45) and was heterogeneous (Supplementary Table 2). Spoligotypes were called correctly for all culture-based samples. Incorrect calls were made for several of the sputum samples, and this is likely due to low coverage of the regions and a consequent miscalling by the Deeplex Web Application.

### Assessment of selected DNA extraction methods in South Africa

DNA extraction protocols that gave promising results in initial testing at UCSF (i.e., bead beating in the Triton X based buffer with a bead cleanup or ethanol precipitation, and heat lysis in the Triton X based buffer with a bead cleanup. The NucleoMAG kit was also done, only done on culture, as the lab in South Africa had experience with this kit) were repeated in South Africa against the reference standard GenoScreen method using clinical samples. All extraction methods yielded sufficient material for Deeplex Myc-TB assay based on Qubit quantification, and we selected three methods for library preparation and sequencing (**Figure 4**). All samples, including those in which drug resistance profiles were indeterminate, had sequencing data of a similar number (2x10^5^ ±7x10^4^; range=7x10^4^, 3x10^5^).

**FIGURE 4:**
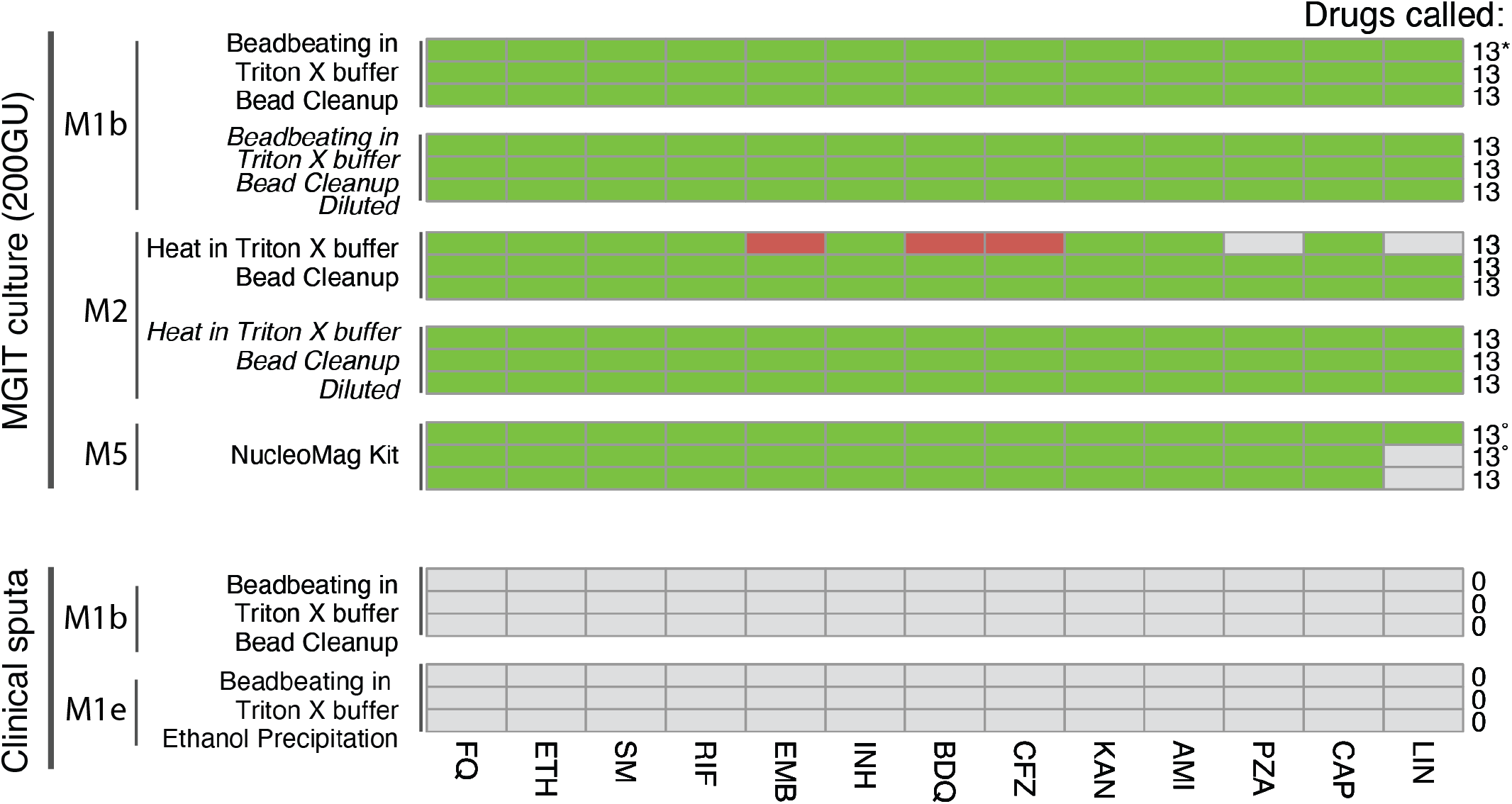
Heatmap of drug resistance call variability from the Deeplex Myc-TB assay done on DNA extracted from clinical sample types using various DNA extraction methods. Since we did not have the known mutations in these clinical isolates, a green block indicates the gene was detected consistently across samples, while red indicates a mismatch for the specific drug (x-axis), and the grey block indicates no or insufficient sequence data, most likely due to no amplification product formed. *Low coverage was reported for all targets, but resistance was still called. °Low coverage was reported for most targets, but resistance was still called. FQ=fluoroquinolones, ETH=ethionamide, SM=streptomycin, RIF=rifampicin, EMB=ethambutol, INH=isoniazid, BDQ=bedaquiline, CFZ=clofazimine, KAN=kanamycin, AMI=amikacin, PZA=pyrazinamide, CAP=capreomycin, LIN=linezolid, MGIT=Mycobacteria Growth Indicator Tube.

#### Sequencing of culture isolates

All methods tested yielded drug resistance calls for most drugs (**Figure 4**). The veracity of resistance calls was unknown due to lack of prior characterization of the isolates.

#### Sequencing of sputum samples

None of the methods tested on sputum produced enough data for resistance calling by the Deeplex Web Application. However, the coverage of the *hsp65* (for *M. tuberculosis* identification) was a median of 10 (IQR 8, 38) and 1 (1, 2) while the number of drug resistance gene targets was 1 (IQR 1, 2) and 0 reads for the bead cleanup and ethanol precipitation methods, respectively.

#### Off-target sequencing

Samples extracted from sputum using bead beating lysis followed by a bead cleanup or ethanol precipitation had only 7.5% (7.1, 9.9) and 5.7% (IQR 5.0, 6.0) alignment to the *M. tuberculosis* H37Rv genome, respectively. Most of the sequenced DNA aligned to the human genome (71%±6%) and likely originated from the sputum samples.

## Discussion

We describe our initial assessment of various methods for extracting *M. tuberculosis* DNA from early MGIT culture-positive samples and directly from patient sputum for downstream use in the Deeplex Myc-TB targeted NGS assay to provide insights to laboratorians newly setting up these workflows intended for routine laboratory settings. We compared several cell lysis and DNA purification methods, including bead-beating and heat lysis with ethanol precipitation or magnetic bead cleanup, and the manufacturer-recommended user manual method (which uses bead beating and ethanol precipitation). To demonstrate that different assays or intended endpoints require different workflow evaluations, and to emphasize the need for methods to be validated with the specific assay output in mind, we evaluated extracted DNA using a multiplex TaqMan-based qPCR and with sequencing the Deeplex Myc-TB prepared amplicon libraries. Except for the NucleoMAG kit method, which did not work at UCSF, all methods gave detectable DNA for all three targets with qPCR, yet they all performed differently in the Deeplex Myc-TB assay. This is likely due to the different robustness of the two reactions, with the Deeplex Myc-TB assay likely using a high-fidelity polymerase which is much less robust than the polymerase used in our qPCR assay.

Bead-beating with magnetic bead cleanup yielded good sequencing coverage for both culture and sputum samples. Moreover, this method also performed well regarding reproducibility, as we observed consistent results across multiple experiments and different batches of samples. However, we did note that diluting the sample produced cleaner results, likely because of lower concentrations of PCR inhibitors. This is likely due to the carryover of PCR inhibitors which are then diluted to a lower concentration. Adding bovine serum albumin (BSA) in the DNA elution buffer may also help alleviate the effect of PCR inhibitors^24^.

For sputum samples, only bead beating with a bead cleanup and the Deeplex Myc-TB manual method produced sequencing data for all targets at the two different bacterial loads; however, all five methods produced similar amounts of total sequencing data, likely due to the substantial amount of human DNA sequenced. This is explainable since the Deeplex Myc-TB assay uses a downstream tagmentation library preparation process; any DNA present in the sample (human DNA being of particular concern within sputum samples) can become part of the library and, moreover, very long fragments of DNA (even at concentrations much too low to see on a gel), would be more favourably tagmented than smaller fragments. Including Illumina adapter sequence on the amplification primers would circumvent library prep and this issue.

Several variations occurred on retesting the methods “in the field” using patient-derived culture and patient sputum samples. This is often the case when performing protocols in different labs, and if not reported, can cause the incorrect interpretation of results. For culture isolates, bead beating and heat lysis performed similarly (both appeared to have PCR inhibitors which were easily diluted out [the volume of culture used was much higher than in the UCSF testing]). Both provided enough data for the high confidence calling of mutations at all targets. At the same time, the NucleoMag kit had a similar number of reads sequenced, but shallow coverage across most of the Deeplex targets (most of the reads mapped to the *M. tuberculosis* genome). For the sputum samples, no method gave data for which the Deeplex Web Application called resistance, and most of the sequence data did not map to the *M. tuberculosis* genome. The presence of large amounts of human and non-mycobacterial bacterial DNA, and the presence of inhibitors may explain this. Including BSA in the elution buffer and increasing the number of PCR cycles may alleviate some of the inhibition. The additional heat steps likely degraded some of the DNA compared to the previous experiments, and there was some sample loss in the bead beating due to the use of snap cap (as opposed to screwcap) tubes in South Africa.

We analysed the data for drug resistant mutation calling using the Deeplex Web Application and the Deeplex Myc-TB V3.0.1 - Extended catalogue. This application automatically processes files and generates reports. We noted several areas for improvement to optimize use in guiding patient care. Although the assay includes internal positive (non-mycobacterial DNA used as control for PCR inhibition) and external positive (*M. bovis*) and negative controls, the Deeplex pipeline does not assess unaligned reads that may indicate contamination or failure of the amplification PCR. In our experiments on DNA extracted directly from sputum, we noted false positive resistance calls presumably due to this issue, despite all controls being valid. We also note that for *tlyA* (and likely *pncA* as well), the absence of an amplicon *tlyA* is reported as a gene deletion and therefore resistance, as long as there are at least a few partial reads that map. This is suboptimal, since it may represent a dropout of the PCR amplicon (possibly combined with “shotgun” sequences during library preparation), as in our case, and not an actual deletion; the latter would need to be shown by detecting the DNA scar.

The Deeplex assay also contains several PCR targets for delineating spoligotypes and lineage-specific SNPs. Despite *M. tuberculosis* H37Rv being a lineage 4 strain, a lineage-specific SNP was only detected in one sample and was heterogeneous. Spoligotypes were called correctly for all culture-based samples, but incorrect calls were made for several of the sputum samples. This scenario is likely due to low coverage of the regions causing a miscalling by the Deeplex Web Application. We recommend enhanced error handling and reporting in cases where sequencing coverage of known *M. tuberculosis* targets is poor.

The method outlined in the Deeplex User Manual yielded good results. However, this method is lengthy, has many steps, has points where technical variation can easily occur (e.g., ethanol precipitation of DNA and washing of the DNA pellet), and cannot be automated as it requires numerous centrifugation steps, liquid movements, and removal of supernatant without disturbing pelleted material. Automated liquid-handling robots can perform automated DNA extraction, PCR amplification, and library preparation at high throughput. While these systems theoretically reduce lab error and technical variability when employed for the PCR and library preparation assays described here, they are expensive, technically sophisticated to program and maintain, and cannot currently perform *M. tuberculosis* DNA extraction. Pragmatically speaking, the lack of affordable “plug and play” sample processing and DNA extraction systems are a major barrier to scaling up sequencing in high-burden settings. Optimized, simple manual methods that can be fed into automated systems are the current best option for programmatic implementation, clinical utility, and reinforcement of local capacity. Thus, based on the early experience described here, we recommend the bead-beating lysis in combination with DNA capture beads cleanup for optimal extraction of high-quality *M. tuberculosis* DNA for downstream molecular assays, such as the Deeplex Myc-TB targeted NGS assay. This method requires few steps and presents minimal opportunities for technical variability. A bead beater is required, of which there are several (including handheld) variations, though the acceptable speed range has yet to be defined. The method is also easily automatable following the bead beating step, with several already available devices supporting the workflow. Our findings also highlighted the need for standardized protocols for DNA extraction from *M. tuberculosis* to ensure consistency and comparability of results across laboratories.

There were several limitations to this study. First, while our experiments were done in triplicate, we only performed one set of experiments. With the exception of GenoLyse, all methods tested at both sites produced adequate sequencing data on culture. Second, we noted incorrect or inconsistent identification of mutations across replicates within the Deeplex results. However, our goal was to evaluate the DNA extraction method rather than the Deeplex assay itself, its mutation calling pipeline, or Illumina sequencing. It is unlikely that the DNA extraction method contributed to these incorrect or inconsistent calls, as any carryover of chemicals from the extraction method would affect the entirety of the reaction and therefore have produced more misidentified mutation calls across the triplicate assays. Third, the generalizability of the practical aspects of some of our findings, such as the high amount of human DNA being sequenced in sputum samples, should be replicated and clarified. A robust amplification would likely lessen this, but so too could a dilution of the input DNA, better lysis of the host cells, or size selection following PCR to remove large fragments.

In conclusion, bead-beating with DNA capture bead cleanup should be considered for *M. tuberculosis* DNA extraction in NGS workflows in routine laboratories. This method is rapid and limits technical variation, and standardization across laboratories could advance the reliability and comparability of results, paving the way for developing and implementing newer-generation assays for drug-resistant TB diagnosis and treatment.

## Data Availability

All data produced in the present study are available upon reasonable request to the authors

