## Supplementary Tables for "Initial experiences with *Mycobacterium tuberculosis* DNA extraction for downstream Deeplex Myc-TB targeted deep sequencing in a high burden setting"

### Supplementary material

#### Supplementary Table S1: *M. tuberculosis* DNA extraction methods.

| Sample type | Bacterial load | Liquification and decontamination method | Sample number | Lysis method | Extraction method | Measure of success | Success rate | Ref |
|---|---|---|---|---|---|---|---|---|
| Sputum smear-positive | 1+ to 3+ sputum smear grade | Sputa were stored and transported in 1 volume 96% ethanol at room temperature | 1494 (1+ = 576, 2+ = 257, 3+ = 294) | Enzymatic (Proteinase K digestion) | DNA was extracted using the semi-automated Maxwell 16 FFPE DNA purification kit | Deeplex Myc-TB assay | MTBC was identified in 1258 (84.2%) samples; 73.5% (14277/19422) of the expected phenotypes in the 1494 samples were automatically predicted | [2] |
| Sputum smear-positive | 1+ to 3+ sputum smear grade | Thermo-protection buffer and Sputasol for some samples and 99C for 30m decontamination | 16 sputum samples, 3 lymph node biopsies, and 1 bronchoalveolar lavage specimen | Bead beating | AMPure XP bead cleanup | Nanopore whole genome sequencing | A mean depth of 15 achieved >99% five-fold genome coverage in 9/20 clinical samples | [3] |
| Sputum and MGIT culture | 1+ to 3+ sputum smear grade | Unknown | 32 sputa (scanty/1+ = 9, 2+ = 18, 3+ = 16), and 43 MGIT cultures | Bead beating | DNA was extracted on the Diasorin IXT followed by Agilent SureSelectXT enrichment | Whole genome sequencing | 87.5% of sputum samples with high bacterial load samples generated complete genomes compared to 72.2% with medium and 55.5% with low bacterial loads | [4] |
| Spiked sputum | $10^3$ and $10^4$ bacilli | NaOH-NALC | Seven different labs processing 200 samples | Bead beating | Phenol | 35 cycle *IS6110* PCR and PAGE | All labs performed poorly and up to 80% of negative controls were falsely positive. | [5] |
| | | NaOH-NALC | | Boiling | None | 40 cycle *IS6110* PCR and dot blot | | |
| | | NaOH-NALC | | Bead beating | None | 34 cycle *IS6110* PCR and PAGE | | |
| | | NaOH-SDS | | None | Phenol | 30 cycle *IS6110* PCR and southern blot | | |
| | | NaOH-NALC+GTC | | Heat | Silica particles | 50 cycle *IS6110* PCR and ELISA | | |
| | | NaOH-NALC+GTC | | None | Silica particles | 40 cycle *IS6110* PCR and agarose gel | | |
| | | NaOH-NALC+GTC | | Heat | Silica particles | 50 cycle *IS6110* PCR and agarose gel | | |
| Culture | Unknown | Unknown | 2 | French press and lysis buffer containing proteinase K, SDS, and lysozyme. Then a GTC buffer. | Ethanol precipitation | Southern blot | 2/2 | [6] |
| Culture | ~5-40mg pellets | Unknown | Unknown | Enzymatic only | Ethanol precipitation | Nanodrop | 0ng DNA yield | [7] |
| | | | | Bead beating | Ethanol precipitation | | <1ng DNA yield | |
| | | | | Enzymatic only | Ethanol precipitation | | ~2ng DNA yield | |
| | | | | Bead beating | Ethanol precipitation | | ~16ng DNA yield | |
| | | | | Bead beating | Ethanol precipitation | | ~6ng DNA yield | |
| | | | | Bead beating | Isopropanol precipitation | | ~8ng DNA yield | |
| | | | | Bead beating | Column capture | | ~2ng DNA yield | |
| | | | | Bead beating | Column capture | | <2ng DNA yield | |
| Culture | Unknown | 100°C for 30 minutes and the pellet used for extractions | 10 replicates at 4 dilutions each | Enzymatic | Phenol chloroform isoamyl alcohol | IS6110 PCR and agarose gel | 9/10 undiluted | [8] |
| | | | | Ethanol | Ethanol | | 9/10 undiluted | |
| | | | | Boiling | Chelex 100 and Nonidet P-40 | | 9/10 undiluted | |
| | | | | Heat | Chelex 100 | | 9/10 undiluted | |
| | | | | Heat | Chelex 100 and 70% ethanol | | 10/10 undiluted | |
| | | | | Heat and enzymatic | Chloroform and CTAB | | 9/10 undiluted | |
| Sputum and culture | Smear-negative sputum, culture unknown | Heat | 65 smear-negative sputum and 94 MTBC cultured strains | Heat | GeneLEAD (automated SPRI beads) | Deeplex Myc-TB assay | One hundred forty successful Deeplex Myc-TB results were obtained for 46 clinical samples and 94 strains, a total of 85.4% of which had a Deeplex Myc-TB susceptibility and resistance prediction consistent with phenotypic drug susceptibility testing (DST). | [17] |
| *M. paratuberculosis* spiked milk | $10^1$, $10^2$, $10^3$, $10^4$ and $10^5$ CFU/ml | None | Unknown | Boiling | None | PCR of *IS900* | $10^5$ "sensitivity of detection (CFU/ml)" | [12] |
| | | | | Bead beating and boiling | None | | $10^2$-$10^3$ "sensitivity of detection (CFU/ml)" | |
| | | | | Bead beating | Isopropanol precipitation | | 104-$10^5$ "sensitivity of detection (CFU/ml)" | |
| | | | | Bead beating and boiling | Isopropanol precipitation | | $10^2$-$10^3$ "sensitivity of detection (CFU/ml)" | |

| | | | | | | | | |
|---|---|---|---|---|---|---|---|---|
| | | | | Bead beating and lysis buffer | Isopropanol precipitation | | $10^2$ “sensitivity of detection (CFU/ml)” | |
| | | | | Bead beating and lysis buffer | Isopropanol precipitation | | $10^2$ “sensitivity of detection (CFU/ml)” | |
| | | | | Bead beating and lysis buffer | Isopropanol precipitation | | $10^1$ to $10^2$ “sensitivity of detection (CFU/ml)” | |
| | | | | Freeze thaw, boiling and lysis buffer | Isopropanol precipitation | | $10^4$ “sensitivity of detection (CFU/ml)” | |
| Sputa | Low, medium, high | OMNIgene SPUTUM | 8 low,10 medium, and 10 high | prepIT MAX buffer and heat | Kit based ethanol precipitation | Pyrosequencing results for rifampicin and isoniazid resistance markers | 100% | 10 |
| | | NaOH-NALC | | Bead beating | Unknown | | 0% | |
| Smear-positive sputum and MGIT culture | | 95°C for 2 hours | 40 smear-positive samples and 27 corresponding cultures | Sonication | Ethanol precipitation combined with MolYsis Basic5 kit | Illumina whole genome sequencing | All sequenced direct samples produced ≥1.5 million reads but only 168 predictions for first-line (n = 96) and second-line (n = 72) antibiotic susceptibility were made for the 24/37 (65%) direct samples that had at least 3×depth | 11 |
| MGIT culture | (1ml culture) | Sonication and 30 minutes to 2 hours at 95°C | 170 cultures | Bead beating | Ethanol precipitation | Illumina whole genome sequencing | >1 million reads was achieved for 144/154 (94%) isolates | 15 |
| | | | 40 cultures | Enzymatic (kit) | QIAamp DNA mini kit | | 8/40 | |
| | | | 40 cultures | Enzymatic (kit) | QuickGene DNA tissue kit S | | 23/40 | |
| Culture, and spiked sputum, and clinical sputum | 200ul culture in 1.8ml sputum | NaOH-NALC | 8 replicates of spiked sputum | Boiling in Chelex and ultrasonic bath | None | IS6110 qPCR | The proportion of recovered MTB DNA ranged from 35 to 82%. For the Chelex method performed on clinical isolates, the sensitivity of the IS6110 PCR was 100 and 75%, for smear-positive and smear-negative samples, respectively. | 16 |
| | | | 62 clinical sputum samples | | | | | |
| | | | 8 replicates of spiked sputum | Boiling in GTC buffer, freeze thaw and heat | None | | | |
| | | | | Enzymatic and chemical lysis | None | | | |
| | | | | Enzymatic and chemical lysis variation and heat | None | | | |
| | | | | Chemical and heat | None | | | |
| | | | | NaOH-NALC and heat | None | | | |
| Culture | NA | NA | NA | Enzymatic and chemical lysis | Phenol chloroform isoamyl alcohol | NA | NA | 25 |
| Culture | NA | NA | NA | Enzymatic and chemical lysis | Phenol chloroform isoamyl alcohol | NA | NA | 26 |

NaOH-NALC=N-acetyl-l-cysteine–sodium hydroxide, GTC=guanidium thiocyanate, MGIT=Mycobacteria Growth Indicator Tube

**Supplementary Table S2: Deeplex Myc-TB assay strain typing was unsuccessful in most cases, and errors in spoligotyping were common and varied according to the DNA extraction method.**

| | | Lineage specific SNP typing | | | Spoligotyping | | |
|---|---|---|---|---|---|---|---|
| **Sample Type** | **Sample Name** | *M. bovis* at 6.67% | *M. bovis* BCG | No SNP detected | Correct | Incorrect | Not detected |
| **–** | Positive control | 0 | 1/1 | 0 | 1/1 | 0 | 0 |
| **Culture** | Bead-beating in Triton X buffer with bead cleanup | 0 | 0 | 3/3 | 3/3 | 0 | 0 |
| **Culture** | Bead-beating in Triton X buffer with ethanol precipitation | 0 | 0 | 3/3 | 3/3 | 0 | 0 |
| **Culture** | Heat in TE buffer with bead cleanup | 0 | 0 | 3/3 | 3/3 | 0 | 0 |
| **Culture** | Deeplex method | 0 | 0 | 3/3 | 3/3 | 0 | 0 |
| **Culture** | GenoLyse kit | 0 | 0 | 3/3 | 3/3 | 0 | 0 |
| **Sputum 50k** | Bead-beating in Triton X buffer with bead cleanup | 0 | 0 | 3/3 | 3/3 | 0 | 0 |
| **Sputum 50k** | Heat in TE buffer with bead cleanup | 0 | 0 | 3/3 | 1/3 | 2/3 | 0 |
| **Sputum 50k** | Deeplex method | 0 | 0 | 3/3 | 3/3 | 0 | 0 |
| **Sputum 50k** | GenoLyse kit | 1/3 | 0 | 2/3 | 0 | 3/3 | 0 |
| **Sputum 50k** | GenoLyse kit concentrated | 0 | 0 | 3/3 | 1/3 | 2/3 | 1/3 |
| **Sputum 10k** | Bead-beating in Triton X buffer with bead cleanup | 0 | 0 | 3/3 | 3/3 | 0 | 0 |
| **Sputum 10k** | Heat in TE buffer with bead cleanup | 0 | 0 | 3/3 | 0 | 3/3 | 0 |
| **Sputum 10k** | Deeplex method | 0 | 0 | 3/3 | 3/3 | 0 | 0 |
| **Sputum 10k** | GenoLyse kit | 0 | 0 | 3/3 | 0 | 3/3 | 0 |
| **Sputum 10k** | GenoLyse kit concentrated | 0 | 0 | 3/3 | 3/3 | 0 | 0 |
